# Behaviours and attitudes in response to the COVID-19 pandemic: Insights from a cross-national Facebook survey

**DOI:** 10.1101/2020.05.09.20096388

**Authors:** Daniela Perrotta, André Grow, Francesco Rampazzo, Jorge Cimentada, Emanuele Del Fava, Sofia Gil-Clavel, Emilio Zagheni

**Affiliations:** Max Planck Institute for Demographic Research, Germany; Centre for Population Change, University of Southampton, United Kingdom

## Abstract

In the absence of medical treatment and vaccination, individual behaviours are key to controlling the spread of COVID-19. We developed a rapid response monitoring system through an online survey (the “COVID-19 Health Behavior Survey”). Participant recruitment takes places continuously via Facebook in eight countries (Belgium, France, Germany, Italy, the Netherlands, Spain, the United Kingdom, the United States). The survey collects key information on people’s health, attitudes, behaviours, and social contacts. In this paper, we present results based on a total of 71,612 completed questionnaires, collected between March 13-April 19, 2020. We find sex-specific patterns, as women show higher threat perceptions, lower confidence in the healthcare system, and a higher likelihood of adopting preventive behaviours. Our findings also show higher awareness and concern among older respondents. Finally, we find spatio-temporal heterogeneity in threat perception, confidence in organisations, and adoption of preventive behaviours.

---

The ongoing coronavirus disease 2019 (COVID-19) outbreak started in Wuhan City, China, in December 2019 and quickly spread globally, soon reaching pandemic proportions [1]. By mid-April 2020, the virus had already caused over 1.6 million cases and over 100,000 deaths worldwide [2], placing a substantial burden on national healthcare systems and posing unprecedented challenges for governments and societies. As yet, governmental responses to mitigate the coronavirus epidemic have varied considerably across countries. Non-pharmaceutical interventions, specifically intended to reduce sustained local transmission by reducing contact rates in the general population, have so far ranged from moderate containment measures, such as school closures and cancellations of public gatherings, to drastic measures, such as travel bans and nationwide lockdowns [3]. In Western democracies, individual behaviours, rather than governmental actions, are potentially crucial to control the spread of COVID-19 [4]. Human behaviour is in fact a key factor in shaping the course of epidemics [5]. Individuals may spontaneously modify their behaviours and adopt preventive measures in response to an epidemic when mortality or the perception of risk is high, and this may in turn change the epidemic itself by reducing the likelihood of transmission and infection [6, 7, 8].

However, a key problem is a lack of data to assess people’s behaviour and reactions to epidemics. Decision-making and the evaluation of non-pharmaceutical interventions require specific, reliable, and timely data not only about infections, but also about human behaviour. Especially in the ongoing COVID-19 pandemic, where medical treatment and vaccination are still only remote options, mitigation and containment mainly rely on massive and rapid adoption of preventive measures [9]. Understanding how the members of different demographic groups perceive the risk, and consequently adopt specific behaviours in response to it, is therefore key to assess the effectiveness of non-pharmaceutical interventions, design more realistic epidemic models, and enable public health agencies to develop optimal control policies to contain the spread of COVID-19.

We seek to narrow this data gap by monitoring individual behaviours and attitudes in response to the COVID-19 pandemic in multiple countries. In March 2020, we launched a cross-national online survey, called the “COVID-19 Health Behavior Survey” (CHBS), to collect timely data on people’s health status, behaviours, close social contacts, and attitudes related to COVID-19. We recruit respondents through advertisement campaigns on Facebook, that we created via the Facebook Ads Manager (FAM). This novel approach to recruiting respondents allows us to combine the flexibility of online surveys for rapid data collection, with the controlled environment of targeted advertisement. This makes it possible to recruit a balanced sample across demographic groups, that is approximately representative of the general population, after applying appropriate post-stratification weights [10, 11, 12]. Other similar online initiatives have emerged recently [13, 14, 15, 16], but to the best of our knowledge, this is the first cross-national study addressing multiple key factors, ranging from individual behaviours and attitudes to health-related indicators to social contact patterns. Moreover, our sampling approach and continued data collection allow us to compare people’s behaviours across countries that are at different stages of the COVID-19 pandemic, and to assess changes in behaviours after pivotal events, such as nationwide lockdowns.

In this paper, we present first results based on survey data collected over the period March 13 to April 19, 2020 in Belgium, France, Germany, Italy, the Netherlands, Spain, the United Kingdom, and the United States. Over this period, a total of 71,612 participants completed the questionnaire. Our goal here is to provide insights into the relation between participants’ demographic characteristics and (i) the threat they perceive COVID-19 to pose to various levels of society, (ii) the confidence they have in the preparedness of different national and international organisations to handle the current crisis, and (iii) the behavioural measures (preventive measures and social distancing measures) they have taken to protect themselves from the coronavirus. From a public health perspective, this information is key to understand the behaviours and attitudes of specific demographic groups in different countries, and to help guide the decision-making process to design adequate policies to contain the spread of COVID-19.

In the following sections, we describe the main characteristics of our sample and present results on behaviours and attitudes we observed in our sample in response to the pandemic. We close with a discussion and an outlook for the next steps in our broader project. The methodological approach as well as the innovative aspects of participant recruitment via Facebook, can be found at the end of the manuscript.

## 1 Results

### 1.1 Participant characteristics

A total of 71,612 participants completed the survey in Belgium (N=6,253), France (N=6,691), Germany (N=12,442), Italy (N=9,741), the Netherlands (N=5,292), Spain (N=7,491), the United Kingdom (N=8,753), and the United States (N=14,949) in the period between March 13, 2020 (calendar week 11) and April 19, 2020 (calendar week 16). As Table 1 shows, participation by week was high in all countries, with a median number of 2,343 respondents per week in Belgium, 1,490 in France, 3,058 in Germany, 1,810 in Italy, 1,759 in the Netherlands, 2,014 in Spain, 1,114 in the United Kingdom, and 2,496 in the United States.

**Table 1.** Characteristics of participants who completed the COVID-19 Health Behavior Survey during the period March 13–April 19, 2020 in Belgium (BE), France (FR), Germany (DE), Italy (IT), Netherlands (NL), Spain (ES), United Kingdom (UK), and United States (US). Unweighted sample.

|  | BE | FR | DE | IT | NL | ES | UK | US |
| --- | --- | --- | --- | --- | --- | --- | --- | --- |
| No. participants | 6,253 | 6,691 | 12,442 | 9,741 | 5,292 | 7,491 | 8,753 | 14,949 |
| Participants per week |  |  |  |  |  |  |  |  |
| Week 11<br>(March 9-15) | - | - | - | 2,016<br>(21%) | - | - | 1,188<br>(14%) | 1,583<br>(11%) |
| Week 12<br>(March 16-22) | - | - | 998<br>(8%) | 1,937<br>(20%) | - | - | 800<br>(9%) | 2,120<br>(14%) |
| Week 13<br>(March 23-29) | - | 1,374<br>(21%) | 1,590<br>(13%) | 1,849<br>(19%) | - | 1,004<br>(13%) | 3,148<br>(36%) | 3,247<br>(22%) |
| Week 14<br>(March 30-April 5) | 807<br>(13%) | 2,356<br>(35%) | 3,417<br>(27%) | 1,771<br>(18%) | 1,790<br>(34%) | 2,458<br>(33%) | 1,772<br>(20%) | 2,873<br>(19%) |
| Week 15<br>(April 6-12) | 3,103<br>(50%) | 1,605<br>(24%) | 3,379<br>(27%) | 1,085<br>(11%) | 1,743<br>(33%) | 2,404<br>(32%) | 1,040<br>(12%) | 3,088<br>(21%) |
| Week 16<br>(April 13-19) | 2,343<br>(37%) | 1,356<br>(20%) | 3,058<br>(25%) | 1,083<br>(11%) | 1,759<br>(33%) | 1,625<br>(22%) | 805<br>(9%) | 2,038<br>(14%) |
| Sex |  |  |  |  |  |  |  |  |
| Female | 4,203<br>(67%) | 4,712<br>(70%) | 7,731<br>(62%) | 6,337<br>(65%) | 3,511<br>(66%) | 5,128<br>(68%) | 5,706<br>(65%) | 9,833<br>(66%) |
| Male | 2,050<br>(33%) | 1,979<br>(30%) | 4,711<br>(38%) | 3,404<br>(35%) | 1,781<br>(34%) | 2,363<br>(32%) | 3,047<br>(35%) | 5,116<br>(34%) |
| Age group |  |  |  |  |  |  |  |  |
| 18-24 | 1,016<br>(16%) | 1,203<br>(18%) | 2,577<br>(21%) | 2,001<br>(21%) | 705<br>(13%) | 561<br>(7%) | 686<br>(8%) | 1,598<br>(11%) |
| 25-44 | 1,868<br>(30%) | 2,086<br>(31%) | 4,786<br>(38%) | 3,949<br>(41%) | 1,264<br>(24%) | 2,743<br>(37%) | 2,131<br>(24%) | 4,051<br>(27%) |
| 45-64 | 2,174<br>(35%) | 2,217<br>(33%) | 3,391<br>(27%) | 2,633<br>(27%) | 2,004<br>(38%) | 3,078<br>(41%) | 3,626<br>(41%) | 5,120<br>(34%) |
| 65+ | 1,195<br>(19%) | 1,185<br>(18%) | 1,688<br>(14%) | 1,158<br>(12%) | 1,319<br>(25%) | 1,109<br>(15%) | 2,310<br>(26%) | 4,180<br>(28%) |
| Education |  |  |  |  |  |  |  |  |
| Primary school<br>or lower | 298<br>(5%) | 127<br>(2%) | 189<br>(2%) | 354<br>(4%) | 215<br>(4%) | 296<br>(4%) | 97<br>(1%) | 22<br>(0%) |
| Secondary school | 2,717<br>(43%) | 1,353<br>(20%) | 7,635<br>(61%) | 4,819<br>(49%) | 3,079<br>(58%) | 1,889<br>(25%) | 3,254<br>(37%) | 5,668<br>(38%) |
| University level | 2,919<br>(47%) | 4,777<br>(71%) | 4,036<br>(32%) | 4,122<br>(42%) | 940<br>(18%) | 4,465<br>(60%) | 4,061<br>(46%) | 8,989<br>(60%) |
| Other | 319<br>(5%) | 434<br>(6%) | 582<br>(5%) | 446<br>(5%) | 1,058<br>(20%) | 841<br>(11%) | 1,341<br>(15%) | 270<br>(2%) |

Table 1 also shows the demographic characteristics of the participants in each country, based on the unweighted sample. The sex ratio is somewhat skewed towards females compared to the overall population, ranging from 62% female in Germany to 70% female in France. In terms of age, older respondents tend to be over-represented, with a median age of 49 years (IQR 32-62) in Belgium, 46 years (IQR 29-61) in France, 41 years (IQR 28-56) in Germany, 39 years (IQR 27-56) in Italy, 55 years (IQR 38-64) in the Netherlands, 49 years (IQR 36-60) in Spain, 56 years (IQR 41-65) in the United Kingdom, and 56 years (IQR 38-65) in the United States. When it comes to education, there is some variation across countries. More specifically, in Belgium (47%), France (71%), Spain (60%), the United Kingdom (46%), and the United States (60%) most respondents attained university-level education, whereas in Germany (61%), Italy (49%), and the Netherlands (58%) most respondents attained secondary-level education.

After applying post-stratification weights, the bias described above is reduced and the sample approximates the shares reported in nationally representative surveys in terms of sex, age, and educational attainment, as shown in Figure S3 in the Supplementary Materials. For more details, see Section 3 of the Supplementary Materials.

### 1.2 The threat perception of COVID-19

Overall, the threat perception of COVID-19 is highest in Italy with a mean value of 0.69, followed by Spain with 0.68, the United Kingdom with 0.67, France with 0.66, Belgium with 0.65, the Netherlands with 0.62, the United States with 0.61, and lastly Germany with 0.55.

In all countries, there is significant variation in the threat that respondents perceive to different levels of society, by sex and age groups. Figure 1A shows the relationship between the threat perceived by male and female respondents for all levels of society and age groups. Here we observe several patterns. First, the perceived threat is significantly higher among women than among men, except for the threat to oneself and to the family among people aged 65 and over. Second, the perception of threat increases sharply from the personal sphere, i.e. oneself and the family, to more distal contexts, i.e. the local community, the country, and, ultimately, the world. Considering specifically the perceived threat to oneself and to the world, the latter is on average 51% greater. Third, younger people perceive lower threat compared to older people, except for the threat to their family. The latter finding is further supported by Figures 1B and C, which show the relation between the perceived threat to oneself and the family (Figure 1B), and between the perceived threat to the country and the world (Figure 1C), contingent on age. The younger age group (i.e. 18-24) perceives a moderately low threat to themselves with a median value of 0.35 (IQR 0.34-0.38), but significantly higher to their family with a median value of 0.53 (IQR 0.52-0.55). By contrast, the older age group (i.e. 65+) perceives a moderately high threat both to themselves and to their family with a median value of 0.57 (IQR 0.53-0.60). On the other hand, the threat posed by COVID-19 at the national and global level is perceived similarly across all age group, while older individuals (45-64 and 65+) generally perceived the threat as higher. For the sake of completeness, we report the perceived threat posed by COVID-19 broken down by country, age, and sex in Figure S5 in the Supplementary Materials.

**Fig. 1.**
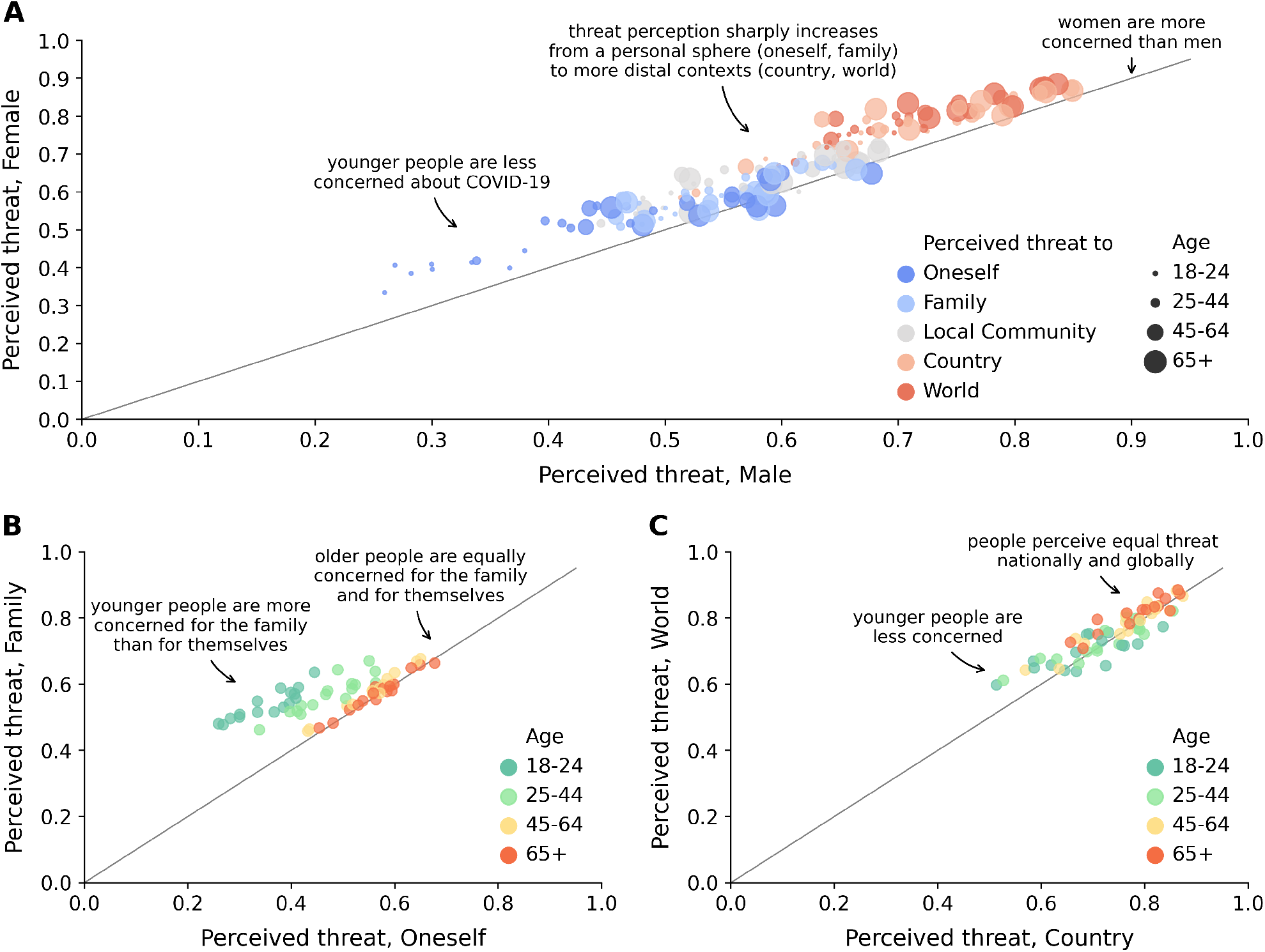
Threat perception of COVID-19. Relationship between the threat perceived by female and male respondents, where colours indicate the different levels of society and sizes indicate the age of respondents (**A**). Relationship between the threat perceived to oneself and to the family (**B**) and to the country and to the world (**C**) by age (indicated by the colour code).

When comparing the threat perception for seasonal influenza with that for COVID-19, we observe that the latter is significantly higher, as shown in Figure 2. In more detail, the perceived threat to oneself is on average 49% higher (ranging from 38% in Germany to 56% in Belgium), the threat to the family is 46% higher (37% in Germany to 52% in Belgium), the threat to the local community is 45% higher (38% in Germany to 54% in Belgium), the threat to the country is 64% higher (50% in Germany to 74% in Spain), and the threat to the world is 54% higher (44% in Italy to 59% in Belgium). More details about the perceived threat posed by influenza can be found in Section 5 of the Supplementary Materials.

**Fig. 2.**
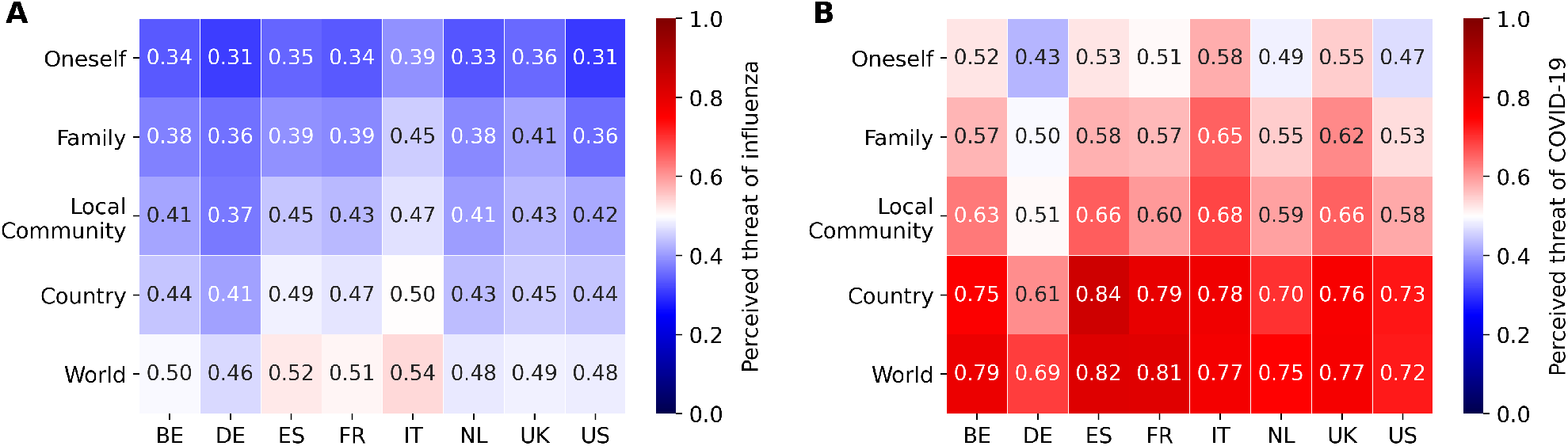
Comparison between the perceived threat posed by influenza and COVID-19. Perceived threat posed by influenza (**A**), and COVID-19 (**B**) to oneself, the family, the local community, the country, and the world. The x-axis reports countries, namely Belgium (BE), France (FR), Germany (DE), Italy (IT), the Netherlands (NL), Spain (ES), the United Kingdom (UK), and the United States (US).

The temporal trend of threat perception differs across countries and it is particularly interesting in Germany, Italy, the United Kingdom, and the United States. Figure 3A shows the weekly percent change of threat perception compared to the initial value in the first week of data collection. In particular, the threat perception significantly varies for Germany, the United Kingdom, and the United States. While in Germany the threat perception slowly decreased over time, in the United Kingdom and in the United States it drastically increased before decreasing to values closer to the ones of week 11.

**Fig. 3.**
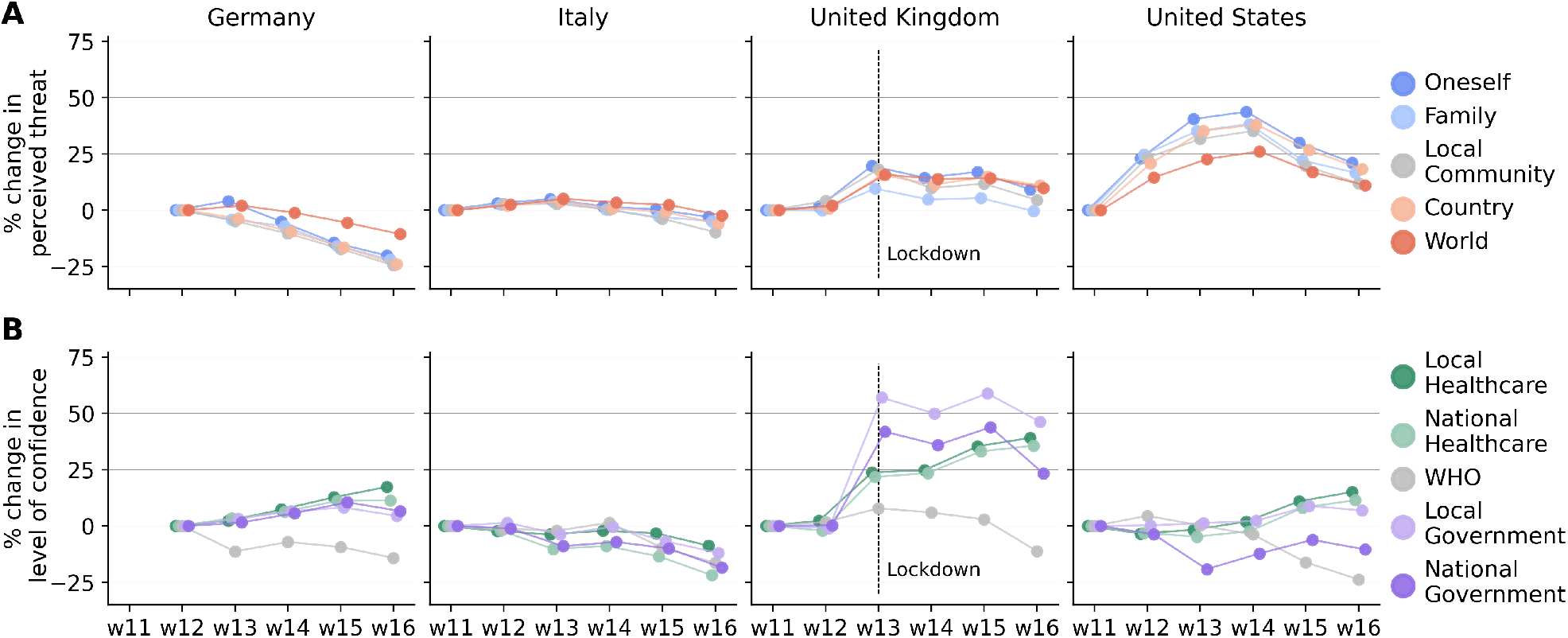
Temporal trend of the threat perception of COVID-19 and confidence in organisations. Weekly percent change in the perceived threat posed by COVID-19 (**A**) and in the level of confidence in organisations (**B**) in Germany, Italy, the United Kingdom, and the United States. The percent change is calculated by considering the initial value in the first week of data collection.

### 1.3 The confidence in organisations

Figure 4A shows the level of confidence that respondents have in the preparedness of organisations to effectively deal with COVID-19. The confidence in the healthcare system is lowest in the United Kindgom and in the United States, and highest in Spain. In particular, people’s confidence in the national healthcare system tends to be lower than their confidence in the local healthcare system, except in Italy and in the United Kingdom, ranging from 2% lower in Spain to 19% lower in France. On the other hand, the confidence in the local and national governments differs substantially across all countries. Confidence in the local and national government is lowest in France, while highest in Germany and the Netherlands, respectively. In particular, people report greater confidence in the national government in Italy, the Netherlands, and the United Kingdom, respectively 6%, 7%, and 13% higher than the confidence in the local government. In the remaining countries, instead, people trust less the national government, ranging from 3% lower in Germany to 28% lower in France.

**Fig. 4.**
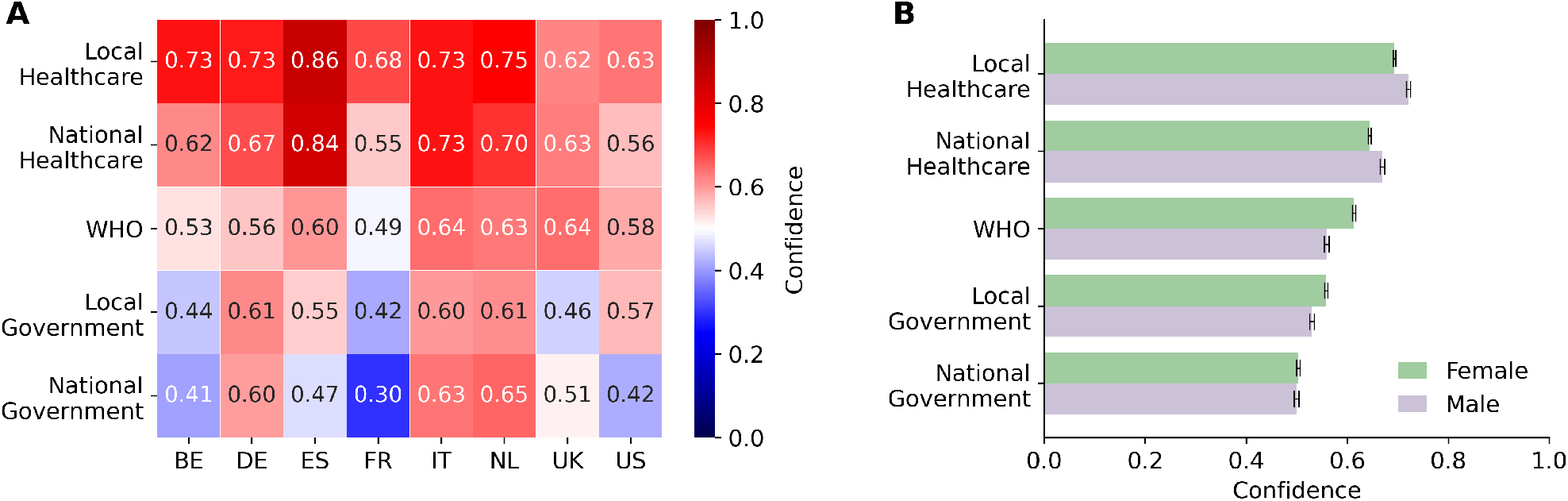
Confidence in organisations to deal with the COVID-19 pandemic. (**A**) Level of confidence in organisations, i.e. the local and national healthcare system, the World Health Organization (WHO), and the local and national government. The x-axis reports countries, namely Belgium (BE), France (FR), Germany (DE), Italy (IT), the Netherlands (NL), Spain (ES), the United Kingdom (UK), and the United States (US). Heatmap shows median values. (**B**) Level of confidence in organisations broken down by sex. Bar plot shows median values and 95%CI as errors.

Examining differences between the sexes in Figure 4B, we observe that on average men tend to be more confident in the local and national health systems, whereas women tend to trust more the WHO and the local government. No significant variation is observed in the confidence in the national government, although there is a substantial difference in the United States, where men have greater confidence in the national government than women. Figure S6 in the Supplementary Materials shows the level of confidence in the organisations, broken down by country, age group, and sex.

When examining the temporal trend, the pattern of confidence in organisations differs across countries and significantly varies for Germany, Italy, the United Kingdom, and the United States. Figure 3B shows the weekly percent change in the level of confidence compared to the initial value in the first week of data collection. In all countries, people lose trust in the WHO over time. In Germany, the confidence in the preparedness of the healthcare systems and governments shows a positive trend. In Italy, people’s confidence slowly decreases, and is down by approximately 16% in week 16 compared to week 11. In the United Kingdom, confidence in the healthcare systems and governments drastically increased after the government decision of locking down the country. On the other hand, in the United States, the temporal pattern is more variable. The level of confidence in the healthcare systems increases from week 15, after remaining constant for nearly a month. We observed fairly stable levels of confidence in the local government, whereas people’s confidence in the national government remained below the initial value.

### 1.4 Preventive behaviours in response to COVID-19

Figure 5A shows the adoption rate of behaviours by country defined as the weighted proportion of individuals who reported having adopted a given preventive behaviour. The least frequent behaviour is the stockpiling of food and/or medicine, ranging from about 18% of individuals in the United Kingdom to about 31% in the United States. Wearing a face mask ranges from about 7% in the Netherlands to about 60% in Italy. As for hand hygiene, the adoption rate of more frequent use of hand sanitizer ranges from about 50% in Germany to about 72% in the United States, whereas the adoption rate of more frequent hand washing ranges from about 87% in Germany to about 94% in Spain. The most frequently reported behaviours are, respectively, the reduced use of transportation, which ranges from about 67% in the Netherlands to about 82% in Spain, and increased social distancing, which ranges from about 93% in the United States to about 98% in Italy.

**Fig. 5.**
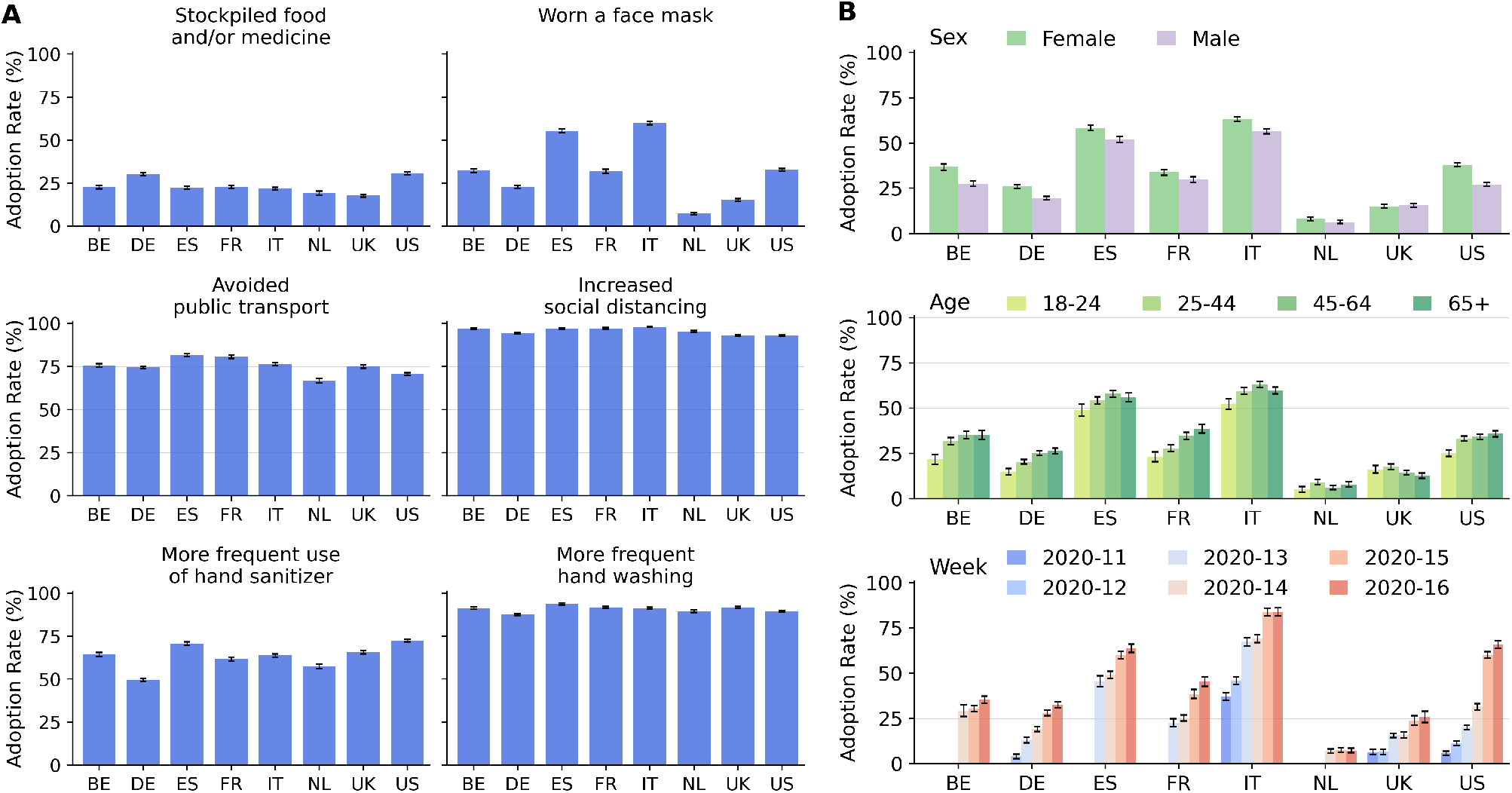
Adoption of preventive behaviours. (**A**) Adoption rate of behaviours by country defined as the weighted proportion of individuals who adopted a specific behaviour. (**B**) Adoption rate of wearing a face mask, by sex (top), age (center), and calendar week (bottom). Bar charts show mean values as bars and 95%CI as errors.

Figure 5B shows the adoption rate of wearing a protective face mask broken down by sex, age, and calendar week. Apart from the Netherlands and the United Kingdom, women and people 45 years or older show the highest adoption rates of face masks. Moreover, the use of a face mask substantially increased over time, except for in Belgium and in the Netherlands. Figure S7B of the Supplementary Materials reports the adoption of behaviours broken down by age, sex, and calendar week. On average, women tend to adopt more protective behaviours compared to men. Social distancing has increased sharply in the United Kingdom and in the United States, whereas it has decreased in Germany, reflecting different stages of the epidemic and different policies.

## 2 Discussion

Understanding how different demographic groups perceive the risk of COVID-19, and thus adopt specific behaviours in response to it, is key to enable public health agencies to develop optimal intervention strategies to contain the spread of the disease. In this paper, we have presented insights from survey data collected through a cross-national online survey, the COVID-19 Health Behavior Survey (CHBS). The survey is ongoing, and here we presented results based on data collected during the period March 13–April 19, 2020 in Belgium, France, Germany, Italy, the Netherlands, Spain, the United Kingdom, and the United States. In this closing section, we summarize the main findings and provide our interpretation in light of the current evidence on the COVID-19 pandemic.

First, we found that the perception of the threat that COVID-19 poses was on average highest in Italy, followed by Spain, the United Kingdom, France, Belgium, the Netherlands, the United States, and Germany. Conversely, respondents’ confidence in the preparedness of local and national organisations to deal with COVID-19 was on average highest in the Netherlands, followed by Italy, Spain, Germany, the United Kingdom, the United States, Belgium, and France. In particular, Italy was the first most affected country in Europe in terms of numbers of cases and deaths, as well as the first country in Europe to implement a nationwide lockdown on March 11, 2020. This may explain the high threat perceived by the population in Italy, and, together with the high confidence in the different health systems and different levels of government, the willingness to adopt preventive behaviours and adhere to social distancing measures.

After Italy, nationwide lockdowns were implemented also in Spain (March 14), France (March 17), Belgium (March 18), Germany (March 22), the Netherlands (March 24), and the United Kingdom (March 24) in order to curb the progression of the virus and to prevent overloading the healthcare system [17]. In the United States, instead, restrictive measures were implemented at the state level, starting in California on March 19, 2020. Note that the lockdowns had different nuances across countries, not encompassing exactly the same restrictions.

Notably, regarding the United Kingdom and the United States, our data collected before and after lockdown measures were implemented (considering the United States as a whole) allow to observe temporal variation in the self-reported behaviours and attitudes: the perceived threat has increased in the population, along with the adoption of social distancing measures. In the case of the United Kingdom, after the lockdown was implemented, the level of confidence in the health systems and different levels of government sharply increased, possibly reflecting discontent in the population about previously announced strategies.

By contrast, in Germany, a somewhat less restrictive lockdown was implemented on March 22, 2020, allowing outdoor activities for families or people living in the same household. Nonetheless, we observe that the adoption rate of social distancing measures was high from the very beginning of our observation period, as well as the confidence in the health systems and both local and national government, which then has further increased over time, contrary to the perceived threat of COVID-19 which instead has decreased over time. This might be interpreted as a case of spontaneous bottom-up behavioural changes emerging from the population, following high trust in decisions and preparedness of the government. Also, of the European countries considered in this study, Germany had the third highest number of cases (about 140,000), but placed only sixth in terms of deaths (about 4,000) as of April 19, 2020 [18], which might explain the lower perceived risk perception in the population.

Second, we observe a clear pattern in threat perceptions regarding different levels of society, sharply increasing from moderate threat for the personal sphere (threat to oneself and the family) to very high threat for more distal contexts (i.e. the local community, the country, and the world). Yet, even though the perception of threat to oneself among our respondents was comparatively low, we found that a high share of them had increased their hand hygiene. This insight renders it uncertain as to what extent behaviour can be straightforwardly linked to perceptions of personal threat. Furthermore, we found that the perceived threat posed by COVID-19 is significantly higher than the perceived threat posed by seasonal influenza. One likely explanation for this is that although seasonal influenza causes regular annual epidemics worldwide [19], the novelty and uncertainty that surround COVID-19 leads risk perception to be substantially higher.

Third, apart from variation at the country level, we found age-specific differences. Looking at the age component, our findings suggest that younger people perceive the threat to themselves lower than older people. This is in line with the evidence that older adults are at highest risk of severe complications following infection from COVID-19 [20]. By contrast, the age structure in the perceived threat to the family is less pronounced, which suggests that respondents were concerned about their family members, regardless of their own age and the perceived threat to themselves.

Fourth, we also found sex-specific patterns in the data. Specifically, female respondents perceived the threat that COVID-19 poses substantially higher, reported a lower confidence in the health system, and were more willing to adopt protective behaviours. Since the case fatality rate for COVID-19 is substantially higher for men [21], we might expect that men are more concerned about it. Our results demonstrate that this is not necessarily true, and that may have to be considered in the design of future communication campaigns.

We gained these insights by using a novel approach for collecting health behaviour data in times of a pandemic. We employed Facebook advertising campaigns to continuously recruit a large number of participants for our survey over a long period of time. This approach allows us to target specific demographic groups in a comparative, cross-national approach, and to collect balanced samples to which post-stratification methods can be applied. To the best of our knowledge, our survey provides the most comprehensive and rigorous cross-country and comparative data on health status, attitudes and behaviors during the peak months of the COVID-19 epidemic in Spring 2020 in Europe and in the United States.

These advantages notwithstanding, our approach also has some limitations. First, online surveys potentially suffer from bias due to self-selection and non-representativeness of the sample. In the case of Facebook, there is increasing evidence that samples obtained from this social media network are not significantly different in central demographic and psychometric characteristics from samples obtained by more traditional recruitment and sampling techniques [12]. Furthermore, by applying post-stratification weighting, which is a standard procedure in survey research, we can correct for non-representativeness in observable characteristics (but not necessarily for self-selection based on unobservable characteristics), at least at the level of the entire sample. Ideally, in our cross-temporal comparisons, we would apply this approach at the level of the week, to warrant complete comparability of observations over time, but issues of data sparsity complicate this approach. We do not expect this to strongly affect our results, but it should be kept in mind that our weekly results might suffer from somewhat larger bias than our aggregate results.

Second, our data collection started at different time points across countries, and also pertains to different points in the trajectory of the pandemic across countries. This also encompasses differences in the implementation of non-pharmaceutical interventions ordered by local and national governments, and needs to be kept in mind when comparing and interpreting our results across countries.

Third, the data presented here have the form of repeated cross sections, which enables us to assess changes in the population samples over time, but does not allow us to assess changes within individuals.

We are planning to address some of the limitations in the future in the following way. First, we aim to expand our post-stratification weighting scheme, by applying multilevel poststratification, which will enable us to achieve greater consistency among differently sized strata and greater precision in the estimates for population subsets, such as the weekly estimates presented here. Second, we aim to carry out a follow-up survey among participants who agreed to provide their email address for this. This panel perspective offers a unique possibility to understand how the COVID-19 pandemic affects the population in the long run and to assess the impact of loosening the lockdown measures on social contact patterns and health behaviours in a cross-national perspective.

To conclude, our work reduces the gap in human behavioural data, by providing timely and accurate data on individual behaviours and attitudes across countries. Our work also illustrates how social media networks, like Facebook, together with appropriate survey designs and statistical methods, offer an innovative and powerful tool for rapid and continuous data collection to monitor trends in behaviours relevant for mitigation strategies of COVID-19. Taken together, the insights gained from our survey data are particularly relevant for policy makers and help design appropriate public health strategies and communication campaigns, and to design realistic epidemic models, which can account not only for the spatio-temporal spread of the infection, but also for accurate data on individual human behaviours.

## 3 Methods

### 3.1 Study design and data collection

The CHBS is designed to collect information on respondents’ health behaviours and attitudes related to COVID-19. Participation in the survey is anonymous and voluntary. Respondents can stop participating at any time and can skip questions they feel uncomfortable answering.

The questionnaire consists of four topical sections: (i) socio-demographic indicators (age, sex, country of birth, country of residence, level of education, household size and composition); (ii) health indicators (underlying medical conditions, flu vaccination status, pregnancy, symptoms experienced in the previous seven days); (iii) opinions and behaviours (perceived threat from COVID-19, level of trust in institutions, level of confidence in sources of information, preventive measures taken, disruptions to daily routine); (iv) social contact data, i.e. the number of interactions that respondents had the day before participating in the survey in different settings (at home, at school, at work, or in other locations). To facilitate validation and warrant comparability with existing surveys, we included standard questions from several sources, such as the European Social Survey (ESS) [22] regarding socio-demographic characteristics, Ipsos [23] regarding opinions on the coronavirus outbreak, and the Polymod project [24] regarding social contacts. Note that we ask respondents about their behaviour and attitudes related to the coronavirus outbreak only if they indicated that they were aware of it. In more detail, we asked respondents how much, if at all, they had seen, read or heard about the coronavirus outbreak, with the answer options “A great deal”, “A fair amount”, “Not very much”, “Nothing at all”, and “Prefer not to answer”. Respondents who indicated that they knew nothing at all, or that they preferred not to answer, were not asked any further questions related to the outbreak.

We created the questionnaire first in English, and then translated it into the different official languages of the countries in our study, with support from professional translators. We considered country-level differences when adjusting the questionnaire for different countries, where applicable (e.g. differences in the educational system). In the online implementation, the questionnaire is available in both English and the national language(s) of the respective country in which respondents are located. The questionnaire was implemented in the online survey tool LimeSurvey (version 3.22.8+20030) and hosted by the Society for Scientific Data Processing (GWDG). The full English questionnaire (as used in the United States) is reported in the Appendix.

The link to the questionnaire is distributed through advertisement campaigns that we created via the FAM. Facebook is currently the largest social media platform, with 2.45 billion monthly active users worldwide as of September 2019 [25]. In the United States, about 69% of adults used Facebook in 2019 [26], with similar penetration rates in Europe, ranging from 56% in Germany to 92% in Denmark [27]. The FAM enables advertisers to create advertising campaigns that can be targeted at specific user groups, as defined by their demographic characteristics (e.g. sex and age) and a set of characteristics that Facebook infers from their behaviour on the network (e.g. interests). An increasing number of studies explore the use of Facebook in demographic and health research to recruit participants for online surveys [28, 29, 30]. Two main advantages of this approach are rooted in Facebook’s wide reach and the possibility to directly target members of different demographic groups. Two concerns that are often raised are that in online samples self-selection might lead to bias in results, and that online sub-populations may not be representative of the general population. However, there is increasing evidence that samples obtained from Facebook do not significantly differ from samples obtained from more traditional recruitment and sampling techniques in central demographic and psychometric characteristics, especially if post-stratification weights are applied adequately [10, 12, 31, 32].

We created one advertising campaign per country and stratified each campaign by sex (male and female), age group (18-24, 25-44, 45-64, and 65+ years), and region of residence (largely following the NUTS classification in Europe and the census regions in US; see Table S4 in the Supplementary Materials), resulting in 24 to 56 strata per country, further stratified using six different ad images. Figure S1 in Section 1 of the Supplementary Materials illustrates the structure of our Facebook advertising campaigns in the United States. Note that we aggregated the different regions of residence into larger macro-regions, to keep the number of strata in Facebook manageable (see Table S4 for the exact mapping). We launched the campaigns on March 13, 2020, in Italy, the United Kingdom, and the United States. We added Germany and France on March 17, Spain on March 19, the Netherlands on April 1, and Belgium on April 4, 2020. In the period 21-26 March, we were unable to recruit a significant number of participants due to technical issues with the FAM which prevented the delivery of our advertisements.

### 3.2 Data pre-processing

We select participants for our analysis in three steps. First, we include only participants who reported that they lived in the country that the respective advertising campaign and country-specific questionnaire targeted, and who reported their sex, age, and region of residence (the central variables in our post-stratification weighting approach, see details below). Second, when analysing responses to a given question, we exclude respondents who chose the options “Don’t know” or “Prefer not to answer”. In the analysis reported here, this particularly affects the question about awareness of the coronavirus outbreak (see Section 3.1); however, as Table S2 in the Supplementary Materials shows, the share of respondents to whom this applies is less than 1% across countries. Third, given that in calendar week 12 we were only able to collect a small number of completed questionnaires in Spain (less than 100), we excluded these data from our analysis as the sample size would render our analysis unreliable for this period. Note that all period references consider local time zones across countries and regions.

After participant selection, we apply post-stratification weights to the final data set in order to correct for potential issues with non-representativeness in our sample. We use a standard procedure in survey research, in which appropriate weights are computed based on population information from more traditional data sources (e.g. census data). Here we use population data from Eurostat (2019) [33] and the US census (2018) [34]. Specifically, for each stratum *i* (given by each combination of sex, age, and macro-region) in each country, we compute the fraction *p*_*i*_ and 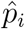 of, respectively, the true population counts *N*_*i*_ and the sample counts 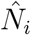 compared to the total population ∑_*i*_ *N*_*i*_ and the total sample size 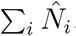. The weights *w*_*i*_ are then defined as 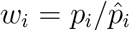, thus giving less weight to groups which are over-represented (*w*_*i*_ < 1) and more weight to groups which are under-represented (*w*_*i*_ > 1) in the sample. We provide more details about our approach to post-stratification in Section 3 of the Supplementary Materials.

### 3.3 Data Analysis

In our analysis, we focus on (i) perceptions of threat from COVID-19, (ii) confidence in the preparedness of different national and international organisations to respond to this threat, and (iii) behavioural measures taken to protect oneself from the virus. All our analyses are based on the weighted sample, whereas the reported sample sizes refer to the unweighted sample.

We asked respondents to rate the threat they perceived COVID-19 to pose for different levels of society (i.e. to themselves, their family, their local community, their country, and the world) on a 5-point Likert-type scale (1 = very low threat, 5 = very high threat), including the options “Don’t know” and “Prefer not to answer”, which are treated as missing values (Table S3 in the Supplementary Materials reports the corresponding sample size for each item). For comparison, we asked respondents to answer the same questions also for the seasonal flu. We normalized respondents’ answers to each item to the range 0-1, meaning that values around 0.5 correspond to moderate perceived threat, whereas 0 and 1 correspond to low and high perceived threat, respectively.

In a similar way, we asked respondents to rate the confidence they had in the preparedness and ability of different organisations to effectively deal with the COVID-19 pandemic (i.e. doctors and healthcare professionals in their community, hospitals in their local area, health care services in their country, the World Health Organization, their local government, and their national government) on a 4-point Likert-type scale (1 = not confident at all, 4 = very confident), also including the options “Don’t know” and “Prefer not to answer”, which are treated as missing values (see Table S3 in the Supplementary Materials). We normalized answers to the range 0-1, and aggregated responses related to the local health system (doctors and healthcare professionals in respondents’ community and hospitals in their local area) by averaging them across items within respondents.

Finally, we asked respondents which measures, if any, they had taken to protect themselves from the coronavirus. For this, we showed a list of actions, from which they could choose all that apply. This list includes preventive measures (e.g. washing hand more often), measures of social distancing (e.g. avoided social events), measures of reduced mobility (e.g. avoided public transportation), panic buying (e.g. stockpiling of food), and potential discriminatory actions (e.g. avoided eating in Asian restaurants). See the questionnaire in the Appendix for the full list of actions. In the analysis, we consider the shares of participants who reported having adopted specific behaviours in response to COVID-19, including: (i) the stockpiling of food and/or medicine; (ii) the use of a face mask; (iii) the increased use of hand sanitizer; (iv) the increased washing of hands; (v) social distancing (if participants selected at least one of the following: avoided shaking hands, avoided social activities, and avoided crowded places); and (vi) the reduced use of transportation (if participants selected at least one of the following: avoided travelling by public transportation, and avoided travelling by taxi).

In our analyses, we used non-parametric tests for median comparisons (Wilcoxon test to compare two groups and Kruskall–Wallis test to compare three or more groups) and considered *p*-values of less than 0.05 to be significant. Data analysis was performed with the programming language Python (version 3.7).

## Data Availability

Due to data protection regulations, the data are not publicly available. For queries about the data, please contact the corresponding authors.

## Acknowledgments

We would like to thank all the participants who took the time to voluntarily complete our survey, and the staff and colleagues of the Max Planck Institute for Demographic Research who contributed to the realization of this project, in particular K. McCann, B. Michaelis, D. Vieregg, S. Leek, C.I. Ruhland, M.J. Bijlsma, M. Lerch, M. R. Nepomuceno, N. Todd, and J. Wilde. This study was funded through the support of the Max Planck Institute for Demographic Research, which is part of the Max Planck Society.

## Ethics statement

This study was conducted in agreement with the data protection regulations valid in Germany. Informed consent was obtained from all participants, enabling the collection, storage, and processing of their answers. All participants’ data was treated anonymously and no personal (and identifiable) information was obtained. Ethical approval for the study was obtained from the Ethics Council of the Max Planck Society.

## Author Contributions

All authors designed the questionnaire and collected the data. DP conceived the project idea, devised the idea for the manuscript, analyzed the data, and wrote the manuscript. AG developed the strategy and technical implementation for data collection and the recruitment of survey participants, and wrote the manuscript. FR supported the strategy development and the technical implementation of the data collection, and wrote the manuscript. JC and EDF designed the post-stratification weighting scheme. DP, AG led the project and the implementation of the online survey. All authors provided input and edited and reviewed the manuscript.

